# Primary Healthcare Innovations in India: Synthesis from a systematic review

**DOI:** 10.1101/2023.07.13.23292645

**Authors:** Angela Chaudhuri, Vijayashree Yellappa, Neha Parikh, Ranjana N Rao, Nilakshi Biswas, Nandini Agarwal, Catherine Cove, Bhumika Nanda

## Abstract

Primary healthcare (PHC) serves as the first point of contact for individuals seeking care. However, the PHC system in India faces significant systemic challenges compounded by multiple disease burdens the population faces. The Astana Declaration highlighted the importance of building a comprehensive and resilient healthcare system, focused on an individual rather than a disease. While Health and Wellness Centers (HWCs) are being developed towards universal health coverage (UHC) as a part of the Ayushman Bharat - Pradhan Mantri Jan Arogya Yojana (AB-PMJAY), several gaps still exist.

A systematic review was conducted following Preferred Reporting Items for Systematic Reviews and Meta-Analyses (PRISMA) guidelines. The study involved developing a theoretical PHC framework, creating search strategies across databases (like MEDLINE, OVID, and CINAHL), and screening them. The review encompassed health innovations and included studies from 1990 to 2019. Relevant quantitative and geographically focused study designs were included, focusing on innovations that improve the efficiency, effectiveness, quality, sustainability, and economy of primary care services.

A total of 239 impact evaluations were included and analyzed. The majority of these evaluations were journal articles (237), with one report and one working paper. The impact evaluations primarily focused on single innovations, although there were also 10 multilayered studies and 7 studies with multiple arms. Out of the 239 innovations, 24 were randomized controlled trials (RCTs) conducted in controlled settings. The studies predominantly took place in rural communities (53%), followed by mixed urban-rural, urban, and tribal communities. Foundations were primary funders (35.6%), with community health worker-delivered interventions, digital service innovations, and supportive mentoring programs being the key supported interventions.

This systematic review offers valuable insights into the challenges and opportunities in India’s PHC system. The findings can inform policymakers, researchers, and healthcare stakeholders in improving primary healthcare delivery and addressing the evolving healthcare landscape in India.

## Introduction

A strong primary healthcare (PHC) system is critical for achieving universal health coverage (UHC) (1,2). The COVID-19 pandemic highlighted the need for resilient primary healthcare systems and revealed fundamental gaps in our approach to primary health. Considering a whole-of-society approach, it is the first level of contact between individuals and healthcare services. It ensures keeping people at the center, taking a comprehensive approach to address socio-economic determinants of health, providing fundamental health interventions and critical preventive activities, and forms the backbone for UHC(3). Further, evidence shows that with an effective PHC system, better health outcomes could be achieved(4,5).

Although access to PHC is expanding globally, around 930 million individuals are still at risk of descending into poverty due to out-of-pocket health expenditures. The WHO predicts that scaling PHC in low-and-middle-income-countries has the potential to save 60 million lives and boost average life expectancy by 3.7 years(6). The discourse around PHC was previously based on curative hospital-centered care, heavily reliant on a biomedical approach. The Astana declaration reinforced the need for a more robust primary health system, moving the focus from single-disease interventions to building a strong comprehensive resilient health system(7). It called upon nations to provide universal healthcare coverage, strengthen primary health systems and encourage people-centered healthcare. This multisectoral approach to health was rooted in social justice, equity, and social determinants of health and focused on mainstreaming the needs of vulnerable communities (8). After the Alma-Ata Declaration in 1978 and the Astana Declaration on PHC in 2018, there was a paradigm shift in the concept of primary health care(9).

### Primary Healthcare System in India

India has identified PHC as an urgent priority. Since independence, India’s health infrastructure has grown with a comprehensive network of about 185,000 government primary healthcare facilities(10).The national task force report on CPHC has categorically mentioned that the PHC is the only affordable and effective path to achieve UHC reiterating the Bhore Committee 1946 recommendations to offer integrated preventive, curative, and rehabilitation services(11,12). However, the PHC facilities are under-function, providing limited care to their population and are riddled with issues of understaffing, infrastructural issues and paucity of medicines and equipment(10). While these issues exist, the National Health Policy (NHP) 2017 acknowledges the need for strengthening primary health systems, recommending that two-thirds of government health spending towards PHC with overall government funding for health to rise to 2.5% of GDP by 2025 (from 1.7% in 2017). In 2018, the Indian government introduced the Ayushman Bharat Program (ABP), which consists of two parts: the Pradhan Mantri Jan Arogya Yojana (PMJAY), which will provide secondary and tertiary hospitalization services to the bottom 40% of Indian families, and Health and Wellness Centers (HWCs), which will strengthen and deliver comprehensive Primary Health Care (CPHC) services to the entire population(13,14). Further, the National Health Policy-2017 has reiterated the need to organize PHC, referral services and collaboration with other sectors(15).

The bottlenecks in the Indian primary healthcare system are common and well documented: shortages in staff, stock outs of common medications, lack of services for NCDs, poor quality of services especially in government PHCs, high cost, and reduced accessibility to other options of private institutions(16). In order to mitigate these challenges and increase access to and strengthen delivery of primary health care, multiple government, NGO, and private initiatives have created comprehensive care models and many have implemented innovative health solutions. In order for its PHC system to address such complex needs of multiple communities and uphold its responsibility of the Alma-Ata Declaration of UHC, it needs innovative solutions that can be flexible and scaled up within low resource settings. However, we cannot have a ‘one-size-fits-all’ approach and there is a need to examine localized PHC innovations that have the potential to strengthen India’s progress towards UHC.

With this background, we undertook a systematic review to document primary healthcare innovations, broadly described as any new or improved intervention, ranging from health policies to management methods to products and technologies. It is meant to respond to any unmet public health need by either creating new resources or streamlining existing ones to focus on vulnerable populations. Health innovations can be created and implemented at any point of the health system and focus on improving efficiency, effectiveness, quality, sustainability, safety, and/or affordability(17). These innovations are crucial to the betterment of CPHCs and filling in the gaps of care to push towards comprehensive care. These solutions are what move the system forward and help program managers and policy makers alike understand new ways to reach UHC.

### Theoretical Framework

We defined primary health care as ‘the first point of contact for patients to help them navigate through changing needs in the life course: addressing health promotion, prevention, treatment, rehabilitation, and palliative care’, meeting the following three components:

1. Meeting people’s health needs through comprehensive promotive, protective, preventive, curative, rehabilitative, and palliative care throughout the life course, strategically prioritizing key health care services aimed at individuals and families through primary care and the population through public health functions as the central elements of integrated health services.
2. Systematically addressing the broader determinants of health (including social, economic, environmental, as well as people’s characteristics and behaviors) through evidence-informed public policies and actions across all sectors; and
3. Empowering individuals, families, and communities to optimize their health, as advocates for policies that promote and protect health and well-being, as co-developers of health and social services, and as self-carers and caregivers to others.

Based on this understanding, we developed a PHC framework and definition to serve as the foundation for the review. We examined several health systems frameworks, including the WHO building block model(18) and adapted it to the vision of a comprehensive, citizen-centric PHC framework indicating how the PHC system’s different components work together for community members (Figure 1).

**Figure 1.**
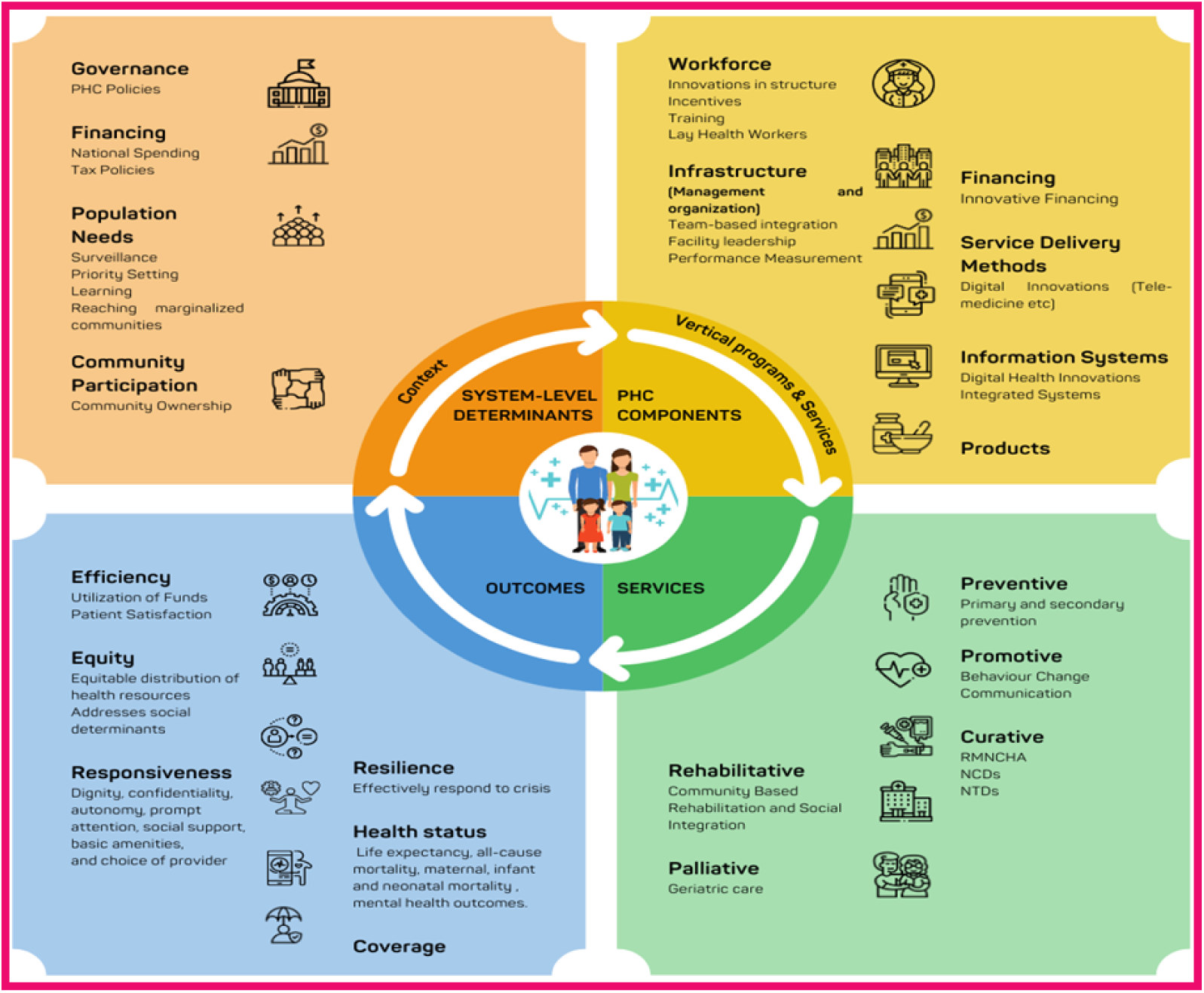
PHC Framework for mapping health system interventions. A Primary Health Care (PHC) Framework was built to solidify the foundation of the systematic review. We considered the WHO building block model, then adapted the framework to incorporate the area of the health system that would make PHC comprehensive, coordinated, and person centered. The final framework to show how all the different parts of the PHC system are mentioned in Fig 1.

## Methodology

This review was conducted according to the ‘Preferred Reporting Items for Systematic Reviews and Meta-Analyses’ (PRISMA) Guidelines to build the primary healthcare innovation landscape. The review entailed (i) building a theoretical PHC framework, (ii) creating search strategies by categories of the theoretical framework, (iii) running the search strategies, (iv) conducting screening at title, abstract, and full-text levels, and finally (v) extracting data from the studies to understand what types of PHC innovations.

### Approach

The search parameters were based on an amalgamation of frameworks, including the PHCPI (19) framework and WHO building blocks. Health innovations in all health system components were explored to create a landscape of diverse homegrown solutions in India. As a result, a person-centered framework (Figure 1) was developed, enabling us to build broader search strategies to include all health innovations. These elements were subsequently deconstructed into keywords linked to our search strategy (Table 1). Two authors independently searched the databases Medline (PubMed), CINHAL (Ebsco), Cochrane Library, Embase, and Google Scholar. We hand-searched the reference lists of all relevant publications retrieved electronically, allowing us to retrieve additional references. We conducted search strategies to ensure our framework would support our PHC definition and tested for additional keywords not identified in our previous search strategies. Limiters were created as prior inclusion and exclusion criteria, allowing us to screen our search hits.

**Table 1.**
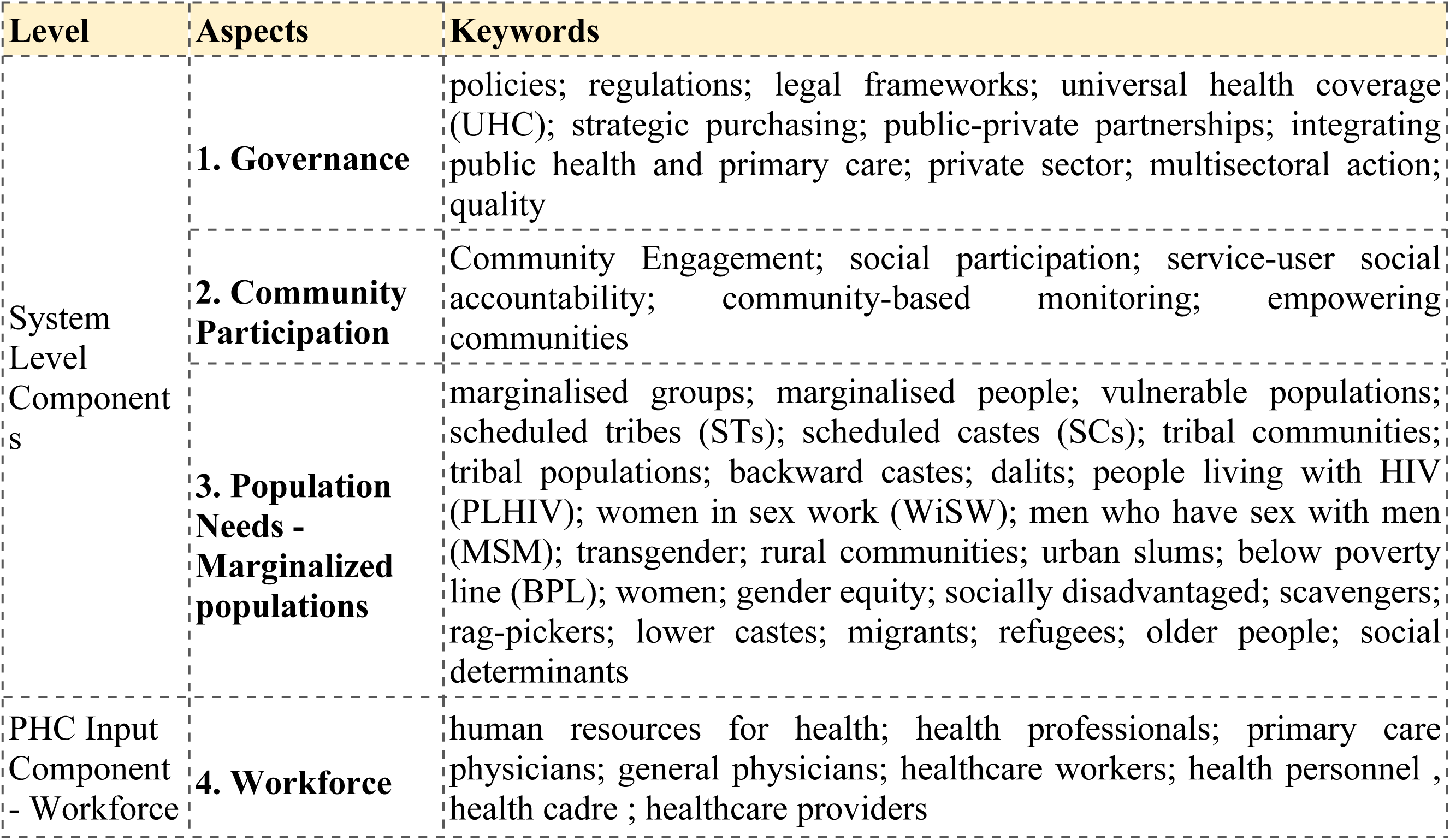
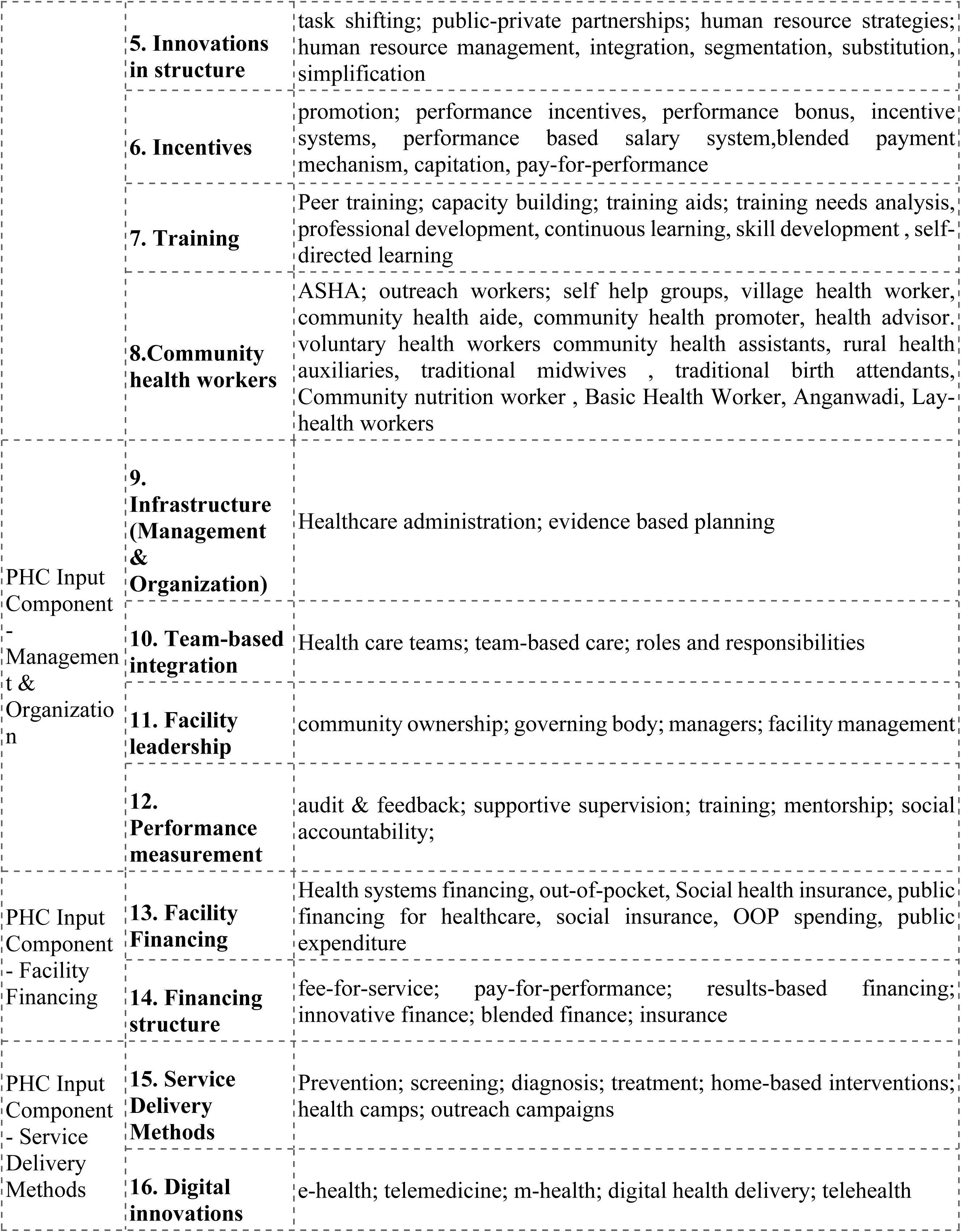
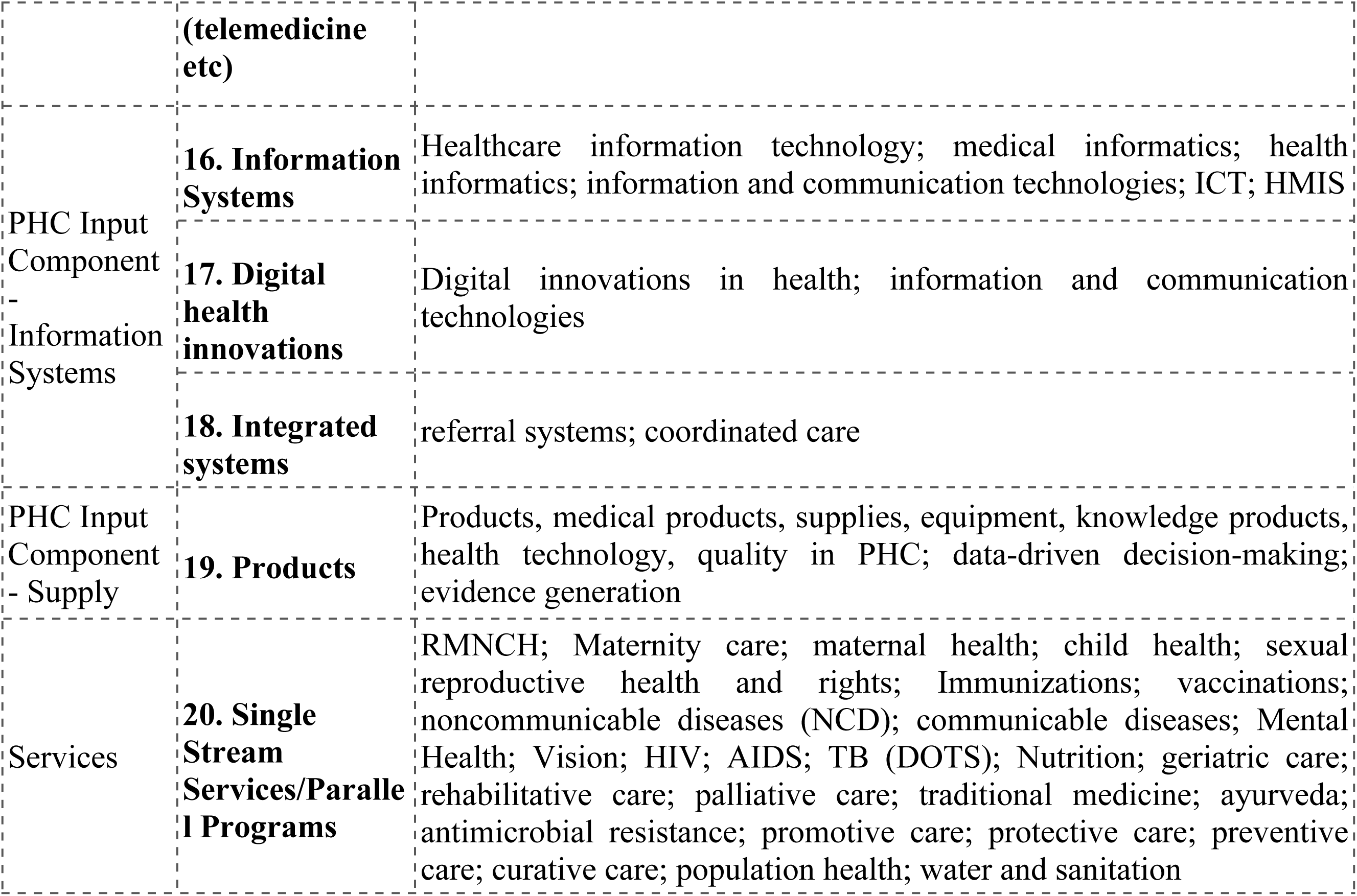
Keyword search strategy.

We did not restrict the year of the search results because this was a comprehensive landscaping review. However, any work before 1990 was not considered as they were no longer relevant to today’s health system. We included studies that highlighted any innovation that enhanced primary care services’ efficiency, effectiveness, quality, sustainability, safety, and economy. This allowed innovations that affected supply and demand, such as performance management studies involving the quality of health services or interventions promoting cervical cancer screenings and health-service utilization. The systematic review included a desk review of peer-reviewed and gray literature (project reports, briefs, working papers and evaluations).

There were also multi-arm and multi-layered studies. Multi-arm studies were defined as studies that may utilize one or more categories of innovations, usually to compare their different effects. For example, an intervention that employs community health workers to screen community members for mental illness and (separately) as another intervention launch a campaign to promote self-care and encourage people to seek care for mental health problems. Thus, this intervention has two arms; one using lay health workers to screen for mental illness and a community participation arm focusing on improving care demand. On the other hand, multilayered innovations layer more than 1 category of innovation in a synergistic way. For example, an innovation that trains an outreach worker to utilize a decision support system and deliver ANC to their respective community would be incorporating both a lay health/outreach worker and a service delivery innovation to reach an outcome of improving infant and maternal mortality. All the multilayered studies had at least two innovations layered within their intervention.

Excluded studies took place outside of India or in secondary and tertiary care settings, as shown in Table 2. These include intervention studies involving medical students or other technical programme students. We also excluded solely observational studies. These comprised studies that documented the prevalence of a disease or behavior or simply described the preferences of a target group without describing innovation or intervention. Finally, in terms of study design, we had no stringent criteria for experimental, quasi, natural, or theory-based interventions. Still, we did exclude clinical trial studies that tested the efficacy of a pharmaceutical drug or product.

**Table 2.**
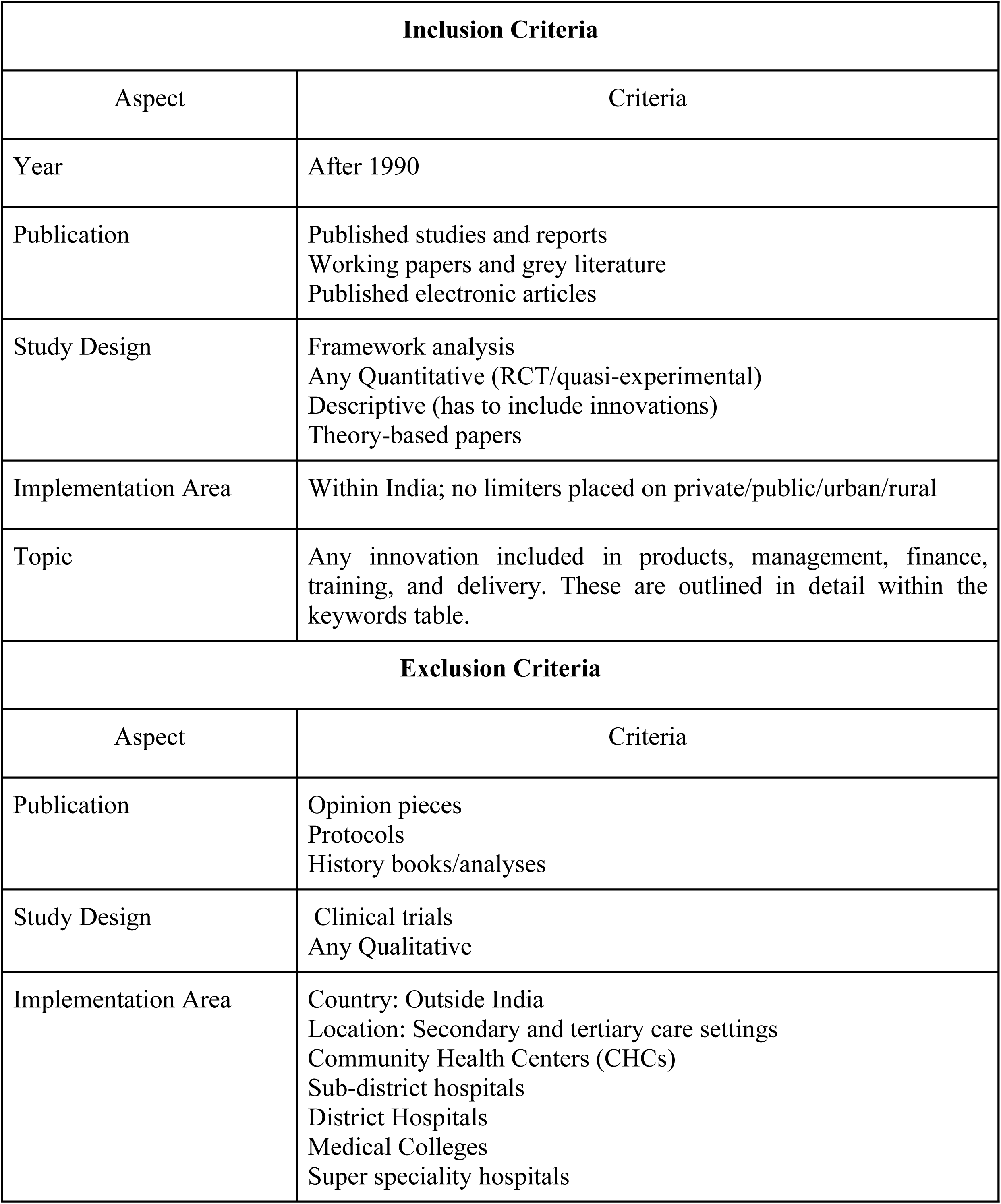
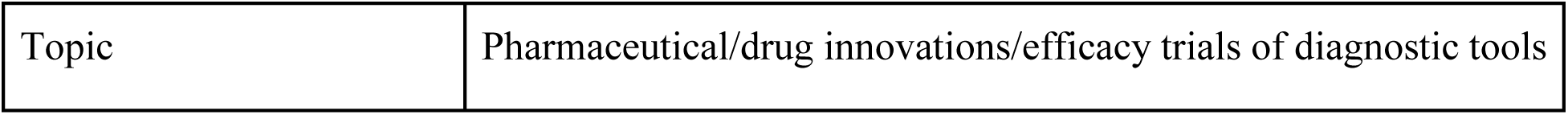
Parameters of PHC Innovations Landscape Literature Review.

We completed our searches and screening through Eppi-reviewer 4, a software created by the University of College London. The uploaded files (7,971.ris files) were then searched for duplicates, including any studies with a 95% match rate. These were then looked through to ensure that a 95% match rate would not delete any linked studies - studies with the same intervention, but published papers documenting different outcomes. Once the first batch of duplicates was deleted, a 90% match rate was used to clean this data again. This resulted in 1,503 duplicates being deleted. Once the duplicates were removed, the subsequent 6,469 studies were screened.

We created codes for screening at the abstract and full-text screening. The ‘include’ and ‘exclude’ codes are given in Table 3. A protocol was created following the order of these codes and the reasoning for selecting each one. In this fashion, any coder could follow the directions on the protocol and complete coding for each study this document.

**Table 3.**
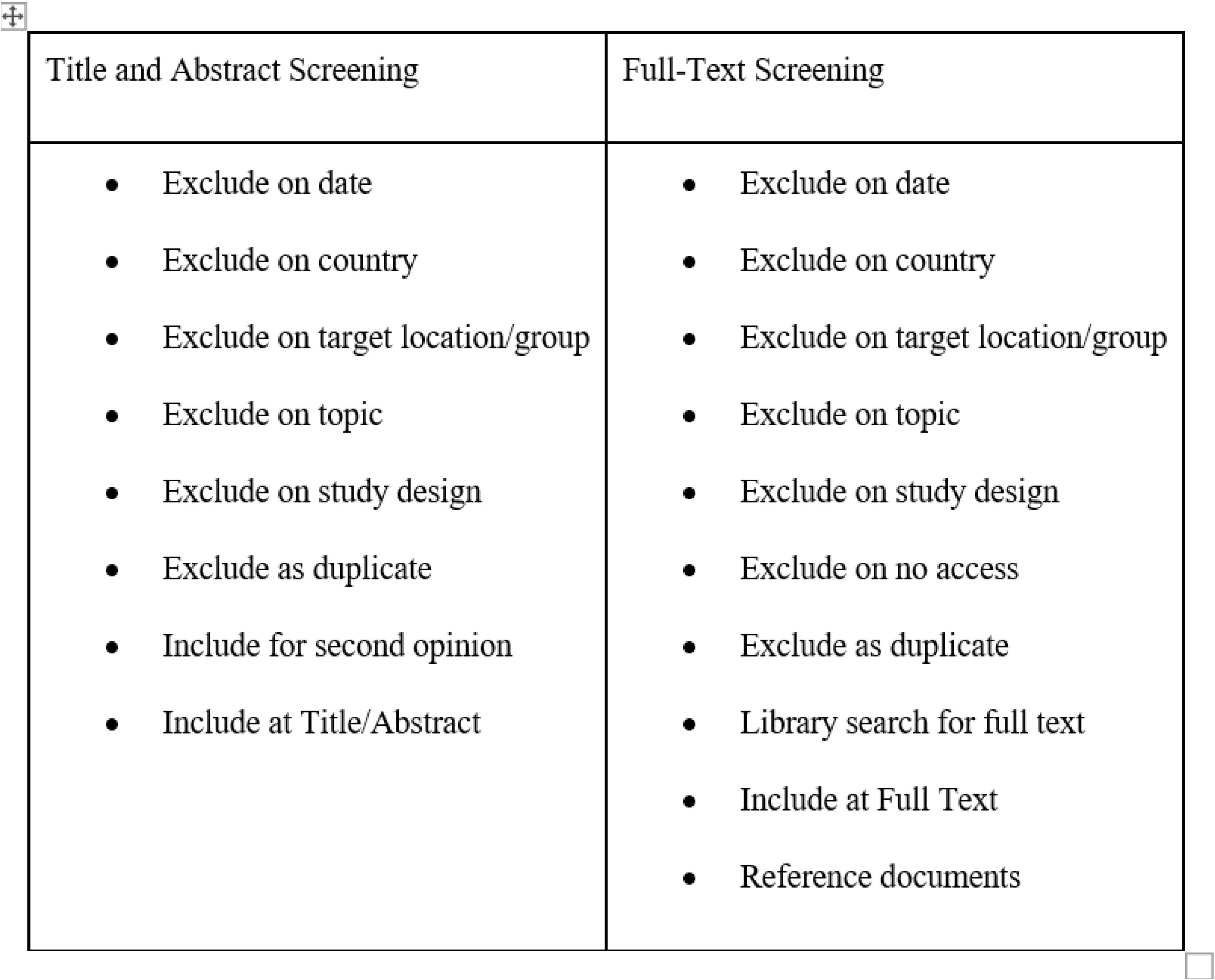
Inclusion/Exclusion Screening Codes.

After screening, feedback was compiled to guide coders in addressing any misunderstandings, inconsistencies, or incorrect coding. This discussion resulted in a greater understanding of our definitions and framework. Coders were given an updated full-text screening process and randomly assigned studies to begin coding. Due to time restrictions, we did not do inter-coder reliability tests or duplicate screening. However, we set up a chat forum where coders could pose questions and come to a consensus on decisions. To complete the project, we added two extra coders throughout the full-text screening phase, and these coders were trained individually. Quality checks were used to eliminate any remaining inconsistencies.

We excluded 4,668 studies and 42 duplicates, the remaining 1,758 studies for full-text screening. 128 studies were excluded on ‘lack of access’. There were 1,333 excluded during the full-text screening, with the majority being excluded by ‘topic,’ followed by study design, setting, country, date, and finally, by no access. This left us with 425 studies to extract and analyze data from. This is represented in Figure 2. 425 studies were then analyzed via an extraction sheet. The extraction sheet included the study’s unique identifier, author, coder, intervention type, study output, funding agency, along with a multitude of demographic data. This extraction sheet allowed the researchers to (i) describe the types of innovations that exist in India and (ii) the type of outputs these innovations hope to achieve. Outputs were categorized as (i)Provider/managerial outputs, (ii)Organizational outputs, (iii)Health and wellbeing outputs (Population level), (iv)Patients outputs, (v) Social outputs and (vi) Unintended outputs. These outputs were designed to encompass the most common goals that PHC interventions hope to achieve. All these outputs lead directly to our PHC framework outcomes of efficiency, equity, responsiveness, resilience, improved health stats, and coverage. Coders were trained on the extraction sheet to ensure they understood the category definitions, how to extract multi-arm and multilayered studies, and how to note any other idiosyncrasies of an uncommon study. With multi-arm or multi-layered studies, each different innovation was coded separately resulting in more documented innovations/extractions than there are publications. Coders were given a total of 3 different publications to read and extract independently and then these were compared in a reliability check meeting, in which extraction details and discrepancies were discussed and finalized via consensus. Through the process of extractions, we excluded an additional 111 studies, some of which were protocols or studies that had interventions but no associated outcome data. This gave us a final number of 314 studies in our final extraction.

**Figure 2:**
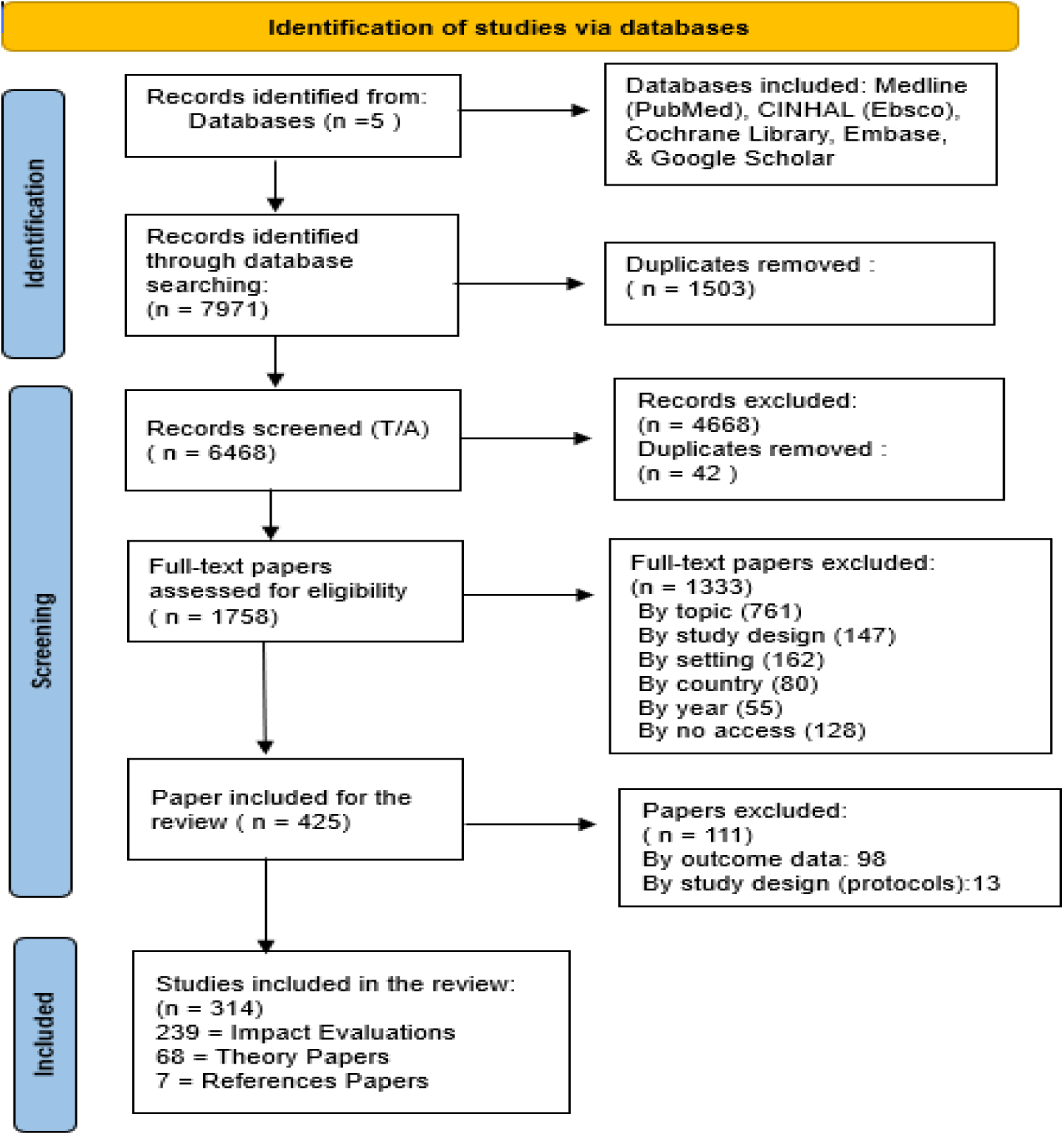
PRISMA flow diagram-. The PRISMA diagram details the search and selection process applied during our systematic review

## Results

In this systematic review, the search yielded 239 impact evaluations that were extracted and analyzed. Although 239 impact evaluation papers were categorized and extracted, 241 innovations were cleaned from this dataset because articles may include many or a combination of innovations documented in our extraction sheet. Most of these impact evaluations were journal articles-237, while 1 was a report and 1 working paper. No editorials or web pages were included in the final dataset.

Most impact evaluations were single innovation-focused studies, with 10 being multilayered and 7 having many arms. Of the 239 innovations, 24 were Randomized Controlled Trials (RCT) in a controlled setting. Most were generally based within rural communities, followed by mixed urban-rural, urban, and tribal communities. It is important to note that there are six times as many studies based in rural areas as in urban ones (53% in rural vs. 12% in urban). Funding provided by foundations (85, 35.6%) catered to lay health worker-delivered interventions (17 of 85, 20%), followed by digital service innovations (9 of 85, 10.6%) and supportive mentoring programs (9 of 85, 10.6%). Most fundings benefited interventions focusing on mobile outreach services (6 of 85, 7.1%) and task shifting among healthcare workers (5 of 85, 5.6%). Academic institutions tended to fund more programs that involved lay health workers (7 of 23, 30.4%). At the same time, funding agencies favored digital service innovations the most (5 of 17, 29.4%). Most government funding went into programs facilitating community ownership (9 of 45, 20%).

### Innovation Category Type

The number of impact evaluations under each category, broad and narrow, are provided in table 4.

**Table 4.**
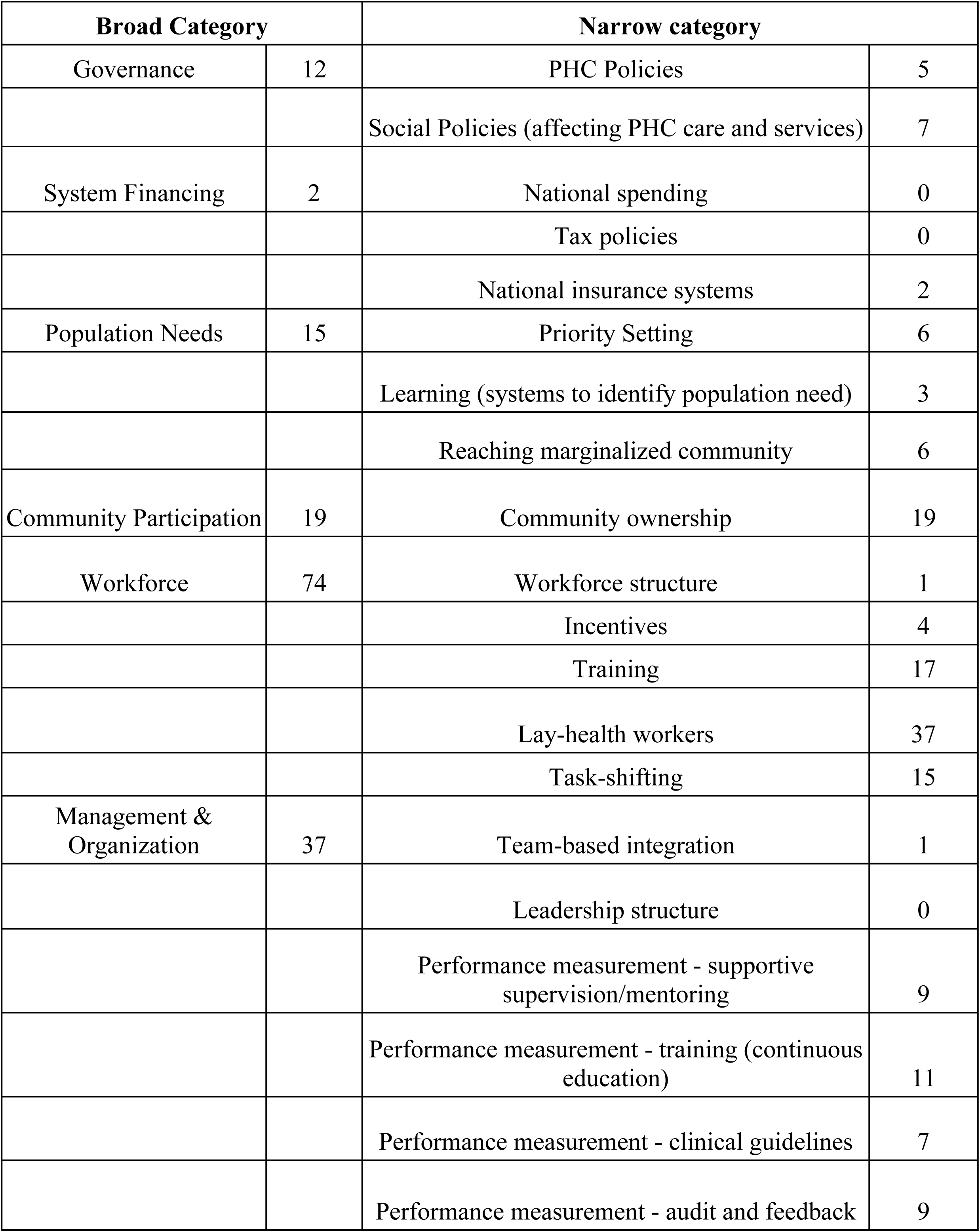
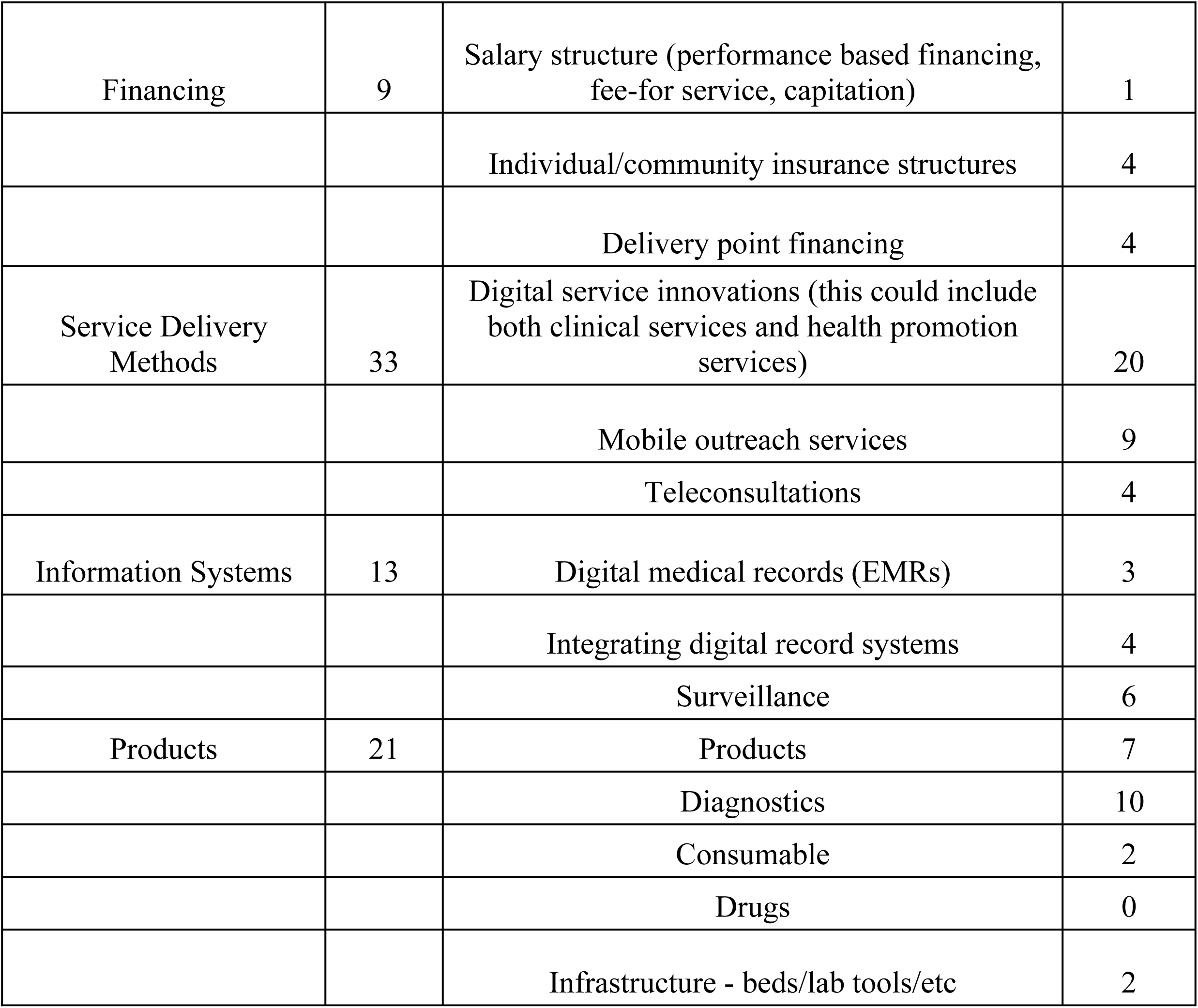
Number of PHC innovations by PHC framework.

The most common areas of emphasis among the 239 innovations were improvements in the workforce (n=74, 31%), management and organization (n=37, 15.1%), and service delivery methods (n=33, 13.8%). The remainder had a similar number of innovations (ranging from 5– 8.8%) save for financing (n=9, 3.8%), which were surprisingly few. There were only 2 systems financing innovations (0.8%), insurance innovations, identified as the lowest among all categories. Employing lay health workers accounted for the majority of workforce category innovations (n=37, 15.1%), with training (n=17, 7.1%) and task shifting (n=15, 6.3%) being the next most widely used type of strategy. Supportive supervision/mentoring, continuous education, clinical guidelines, and audit and feedback innovations (2.9-4.6%) were equally distributed as the innovation type for management and organization. Performance assessment measures, however, dominated this category. None of the innovations under this review addressed the change in the leadership structure. The dearth of innovation to alter the workforce structure (n=1, 0.4%) also supports this. There were various advances in service delivery (n=33, 13.8%). Few used teleconsultations (n=4, 1.7%), and most (n=20, 8.4%) fell under digital service innovation. Multilayer interventions are demonstrated in the table 5

**Table 5:**
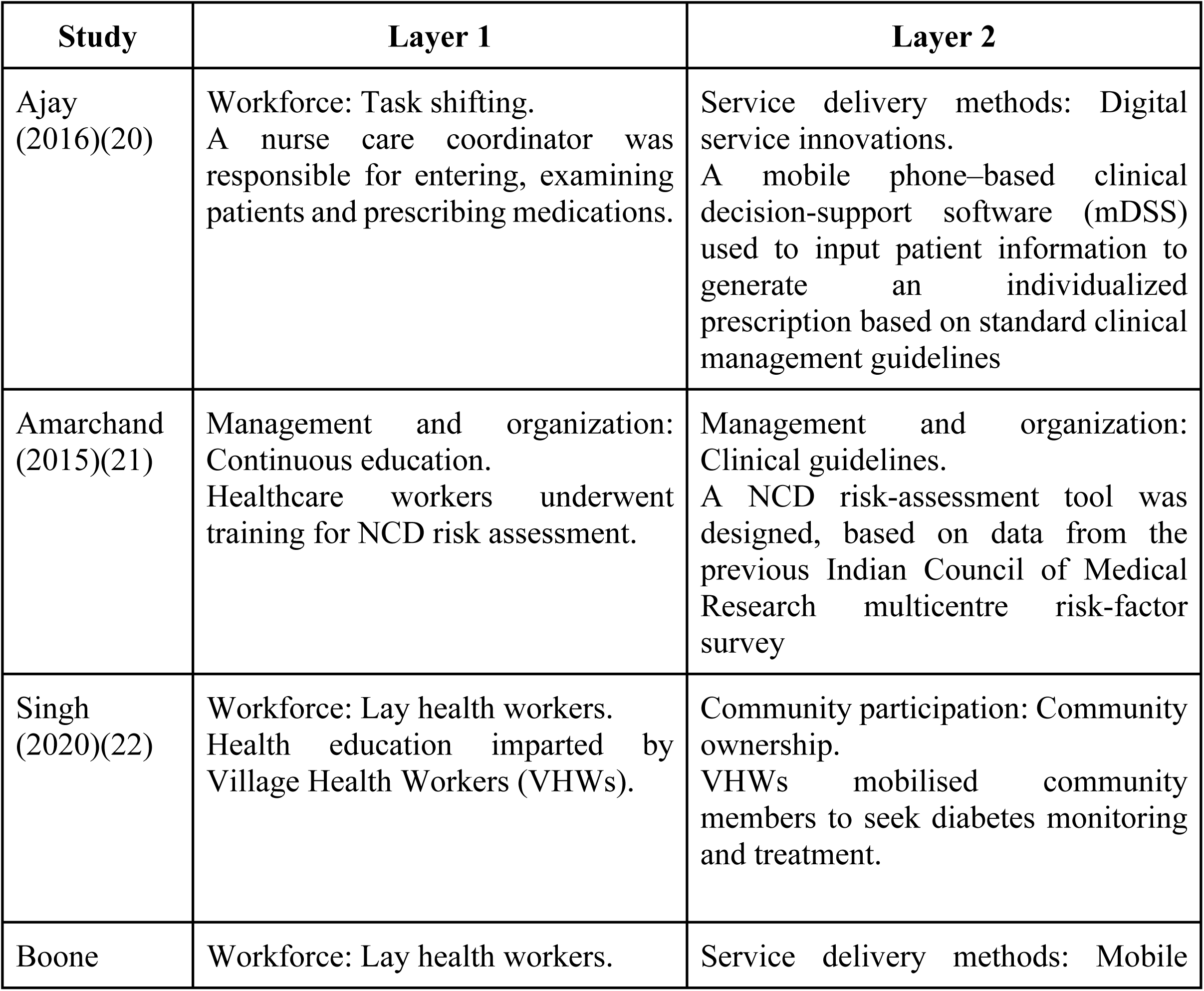
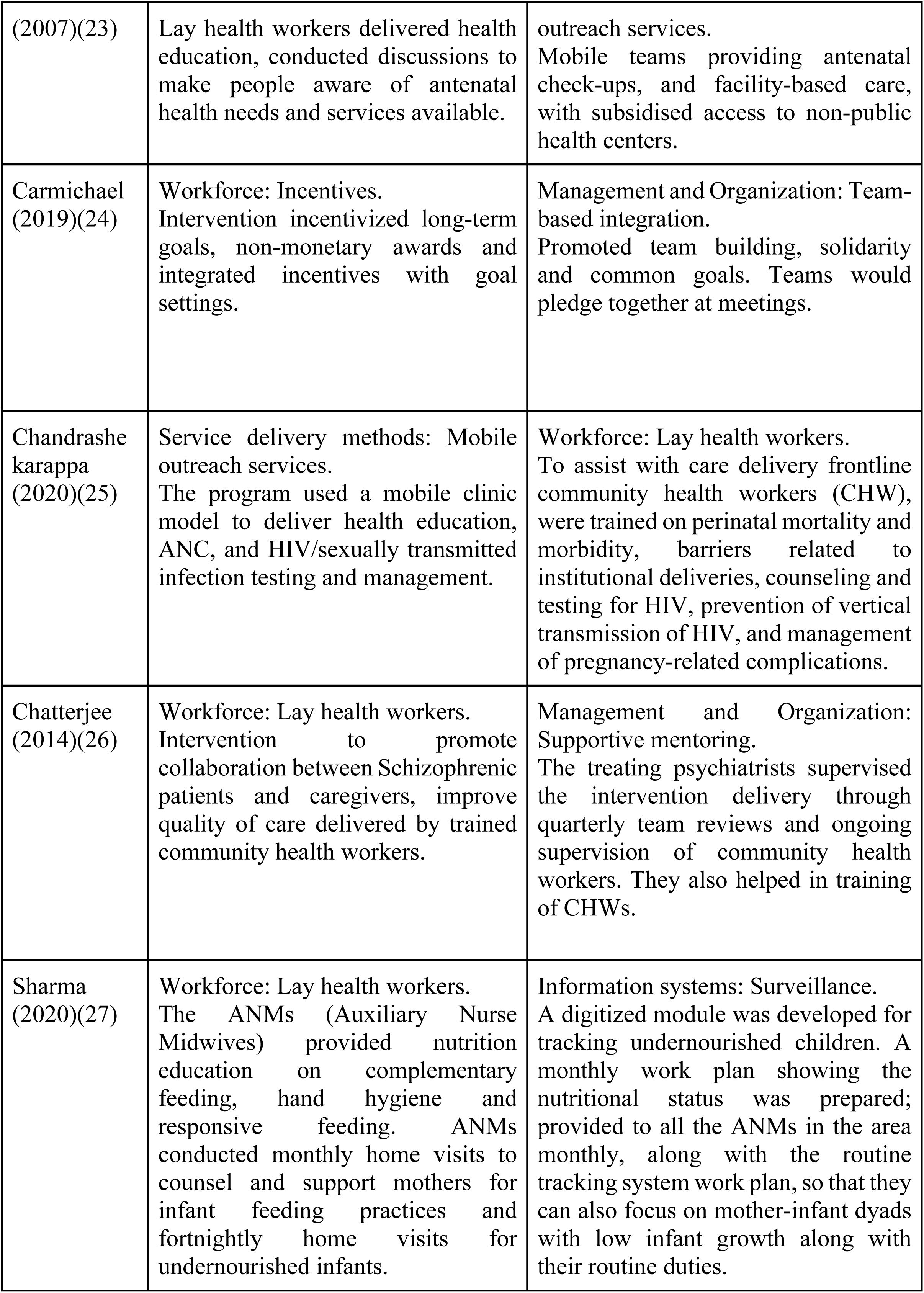
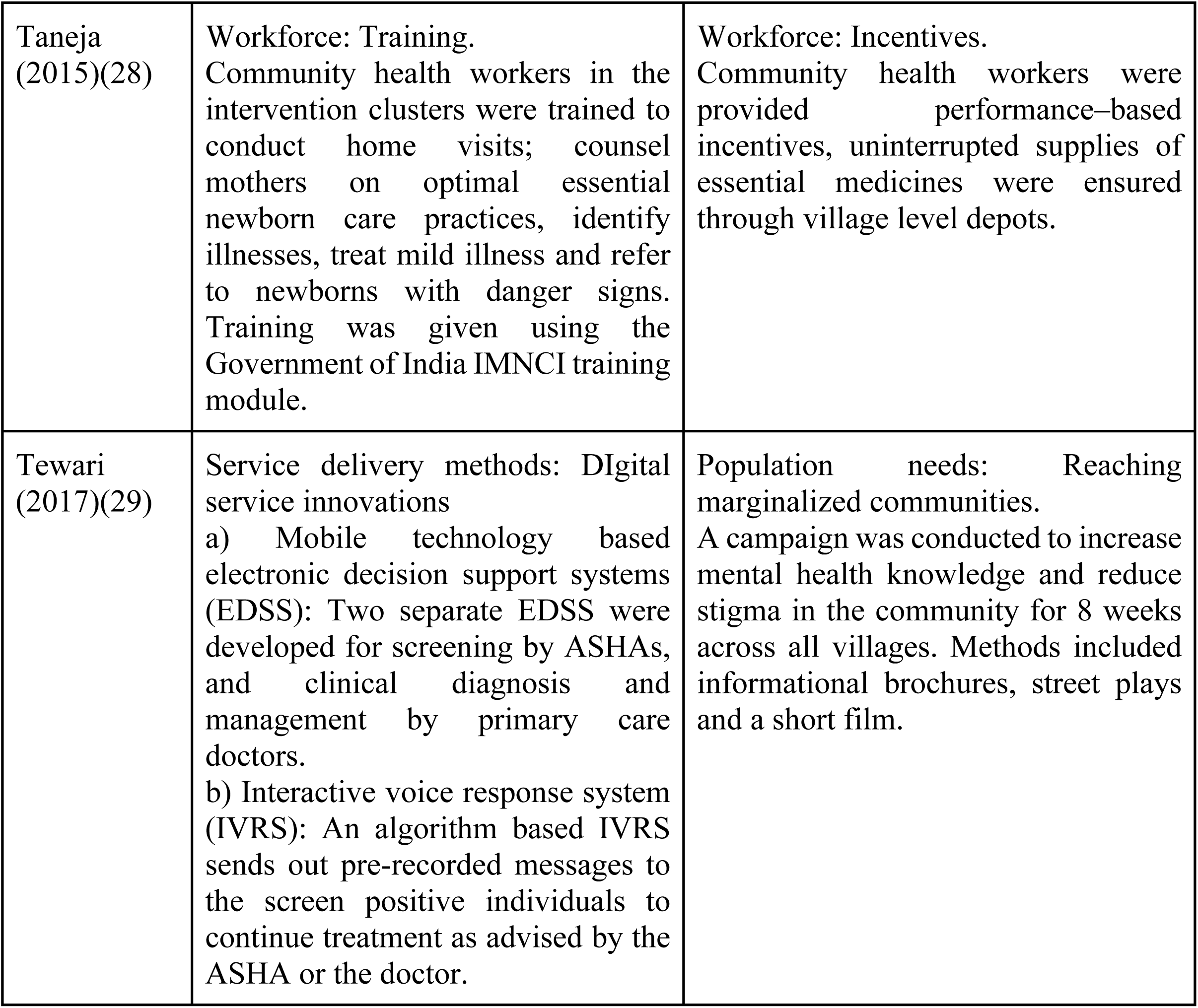
The multilayered studies.

In the following paragraphs, we present the synthesis from systematic review.

### Governance

Primary healthcare governance in India is a complex issue due to the country’s geographical area, diversity, and political structure. The responsibility for healthcare governance in India is shared among the national, state, and local governments and various public and private healthcare providers(30). Interventions that intended to enhance specific components of governance were uncommon. Primary health centers (PHC) policies and social policies are the two subcategories (affecting PHC care and services) in this review. Low socioeconomic households were primarily recipients of these innovations especially in rural areas(31,32).

The government of India launched the National Health Mission to improve the availability, accessibility, and affordability of quality healthcare services, with a particular focus on primary healthcare. The NHM has helped strengthen the governance and management of primary healthcare services in India through various initiatives, including employment schemes(33). In the study on Community Action for Health (CAH) envisioned to strengthen community-based accountability emphasizing the importance of “problematization,” a dimension not currently included in most popular analysis frameworks. CAH highlighted the need for spaces and processes of collective sense-making to ensure that a contested policy intervention was integrated into a complex system (34). A study investigated the impact of India’s National Rural Employment Guarantee Act (NREGA) scheme on the utilization of maternal health services. This study found no significant increase in maternal healthcare utilization, suggesting that income security alone does not reduce barriers to utilization. Additionally, a significant increase in home deliveries was found amongst poorer households, suggesting that Employment Guarantee Schemes (EGS) may introduce unintended barriers that reduce utilization and increase inequity (32).However, advocate for integrated governance potentially improving community participation, service delivery, public system accountability, and human resource rationalization(35). Strategies for innovative institutional structures show improved participation of communities in self-governance, community monitoring of government programs, and, therefore, better services. Integration can affect the decentralization of power, inclusion, efficiency, accountability, and improved service quality in government programs. This was also seen in the study that decentralization of mental health services and integration with general health care is a need for India and is required not only in rural areas but semi-urban areas and suburbs of cities(36).There were detailed institutional structures models provided in the study which can be replicated for the implementation and sustainability of integrated governance (35).

### Information Systems

The innovations under “Information Systems” were relatively evenly distributed across the three subcategories of surveillance, integrating digital records systems and digital medical records (EMRs). While the focus was primarily on rural regions to aid in more efficient data collection and storage, more significant reach in monitoring health indicators including but not limited to pregnancy-related outcomes (37,38) and chronic conditions (39). Most innovations aimed to develop comprehensive and prospective registries to enroll and monitor target populations to facilitate early detection and improved care delivery. These innovations spanned all three subcategories and also came in handy for community health workers and medical professionals. Streamlining data collection and storage and easy access for both provider and patient increased care compliance and reduced loss to follow-up for several innovations (38,40).

The most common targets for interventions to develop or improve existing information systems were population-level health and well-being and provider or managerial outcomes. A few addressed organizational improvements by helping develop data-driven guidelines for disease diagnosis and care management plans (41).

### Products

Health innovations that rolled out some “product” were mainly done in rural regions, focusing on diagnostics and digital products. There was a clear gap in innovations that aimed to improve health infrastructure. Only one representative sample of innovations tested the effectiveness of providing a consumable product (vitamin A supplements) in a school setting(42). Studies also featured cost-benefit analyses for the innovation being tested to determine the product’s feasibility, scalability, and sustainability(43). Digital devices were also standard for early screening, efficient diagnosis, monitoring health indicators, and ensuring compliance with designed care plans(44,45).

Maternal and child health was the primary target for most innovations; however, diagnostic innovations catered to various vulnerable populations. The output was primarily the health and well-being of the community. Some measured organizational outcomes that resulted in more patients receiving care, the feasibility of the product, and sustainability(43).

### Service delivery methods

Service delivery methods were divided into three sub-categories: digital service innovations, mobile outreach programs, and teleconsultations. Innovations in this category were defined as those that improved the effectiveness or efficiency of delivering a health service to an individual. Recent years have seen a spike in digital service innovations, being both a product or an intervention, and a method to spread awareness (20,24,46–52).Especially in rural and tribal regions, digital innovations have taken the lead. Teleconsultations are not as popular but seem to be gaining some traction(53,54). Mobile outreach programs have been the most effective way to reach rural and tribal regions in the past and continue to remain popular(24). However, innovations that have some elements of digital service providers are gaining funding and attention. Considering how widespread cell communication and internet coverage is, reaching several rural and semi-urban populations, it is not surprising that digital innovations are slowly taking the lead. The utilization of community outreach workers such as ASHA workers, Auxiliary Nurse Midwives (ANMs), and social workers was common in outreach and digital innovations.

Several innovations explored provider-related outcomes, such as improved communication within the team and with patients, conducting comprehensive evaluations of the health center, screening, and early diagnosis efforts, feasibility of incorporating digital innovations, and satisfaction of care. Health and wellbeing outcomes were also given priority, and various factors such as mortality, morbidity, hospitalization rates, awareness among people, and satisfaction with quality of care and relationship with providers were measured.

### Financing

Innovative financing was not a very sought-after category, with only two interventions that employed delivery point financing and one showcasing a community insurance scheme making up the representative sample (55–58). Although health financing is an extremely broad category, we searched for financing schemes on both systems and facilities levels that improved coverage and financial safety for individuals whenever they wanted to access health services and financial partnerships that improved healthcare access such as PPPs (public private partnerships). The impact of medical insurance, public-private partnerships and clinics that prioritize sustainability was assessed. Urban regions were the most common regions for these innovations. The primary beneficiaries of all the schemes were people in the lower socioeconomic status with significant burden being put on higher income strata or the government (55,56).However, all the innovations demonstrated a significantly moderate amount of success rate on ability to seek care, hospitalizations, mortality and morbidity.

Outcome types were either organizational or health and wellbeing; measuring care seeking behaviour, health indicators and even early hospitalizations and reduction in adverse events (55–58).The low number of financing interventions in selected clusters are reflective of the general trend seen in terms of primary health interventions.

### Management and Organization

Management and organizational innovations targeted aspects that affect performance of frontline workers, medical professionals and others involved in the care provision process. Audit and feedback, continuous education and training, supportive mentoring and supervision, and clinical guidelines were the subcategories encompassed under the management and organization umbrella. The innovations were fairly evenly divided across the four categories. These interventions usually focus on capacity building and improving workers’ abilities and skill sets(21,59–64).The aim of most of these interventions are early diagnosis, greater vaccine coverage, knowledge of workers, workflow improvement and increased efficiency(65–67). Additionally, performance improvements interventions were very widespread and spanned across urban, rural and tribal regions.

Most interventions that aimed to tackle management issues measured improvement in provider and organizational outcomes. As a result, the effect of such interventions was seen on patients’ health and wellbeing. Mentoring and feedback based innovations were seen gaining popularity in recent years(64–66), while training and continuous education programs have always been prevalent.

### Workforce

Workforce development has been the most targeted of primary health interventions in the past and continues to remain popular. Workforce interventions were any type of intervention that affected change in Human Resource for Health (HRH) that resulted in improved healthcare delivery including the following subcategories: workforce structure, training, incentives, lay-health workers and task-shifting. Lay health workforce development, training, capacity building and utilization was the most common type of workforce innovation. Task-shifting, followed by training and incentives were also seen throughout. Workforce structure was the least common type of innovation with not one in the selected clusters. The current network of ASHA workers and ANMs (Auxiliary Nurse Midwives) has proven to be highly effective for several types of innovations, such as vaccination programs, screening and early diagnosis, care monitoring and health management, and health education and awareness campaigns(22,68–74). Such interventions were equally divided into rural and urban regions.

Workforce interventions attempted to improve the large-scale health and well-being of target populations. They measured health indicators, help-seeking behavior and practices, knowledge levels, and reduction in morbidity and mortality rates, especially in maternal and neonatal health(71,72,74–77).Organizational and provider impacts were seen in improved care delivery, reduced medication over-prescription, increased successful engagement with care, and low refusal rate for screening programs (68,72,75,78,79).

### Community Participation

Community participation interventions focussed on promoting community ownership. The involvement of the target population in their own care and treatment and the decision-making process was critical to these interventions. These interventions were implemented in rural regions and employed the tight-knit community dynamic to ensure success and sustainability(80–82).All the selected programs saw empowering the people and equipping them with the knowledge and skills to take charge of their care. Such interventions have been seen increasingly in recent years. Health and well-being of the community and individual health indicators were primary outcomes in all these programs. Reduction in morbidity and mortality was seen due to improved knowledge and practices by the people (80–82).

### Population needs

Population needs interventions were those that didn’t focus on individual care delivery, but rather systemic improvements in the health system that would affect large communities and populations. Its subcategories included surveillance and reaching marginalized populations. These innovations were not as common and were irregularly seen in the last decade (83,84). Surveillance interventions mainly focused on screening and early diagnosis of communicable and non-communicable diseases (83).Whereas interventions that aimed to reach marginalized populations tackled social inequities affecting health (84). All such interventions were implemented in rural regions. However, they were not very common and weren’t seen as much in the latter part of the last decade.

Impact on the health and well-being of the population was the primary outcome of all of these interventions. The goals ranged from mapping cases with specific diseases or conditions, studying the impact of specific treatment, and targeting inequities and problems in marginalized communities(83,84).

### Systems Financing

System financing interventions were divided into three subcategories: national spending, tax policies, and national insurance schemes. This was the least common type of health innovation seen, with the only included intervention reporting on a national insurance scheme, Rashtriya Swasthya Bima Yojna (85). This national scheme was evaluated for its impact on beneficiary households’ utilization of health services, per capita out-of-pocket (OOP) expenditure, and per-patient OOP expenditure on major morbidities(85). Large gaps exist in the field of financing and are reflected by the lack of suitable interventions in the selected clusters.

### Intervention Outputs

Health outputs involved changes in care utilisation, improvement in care delivery on a local and national level, and improved provider and organizational accountability. Maternal and child health schemes were prioritized over other issues and health problems (31,86,87).

Most interventions looked at either a combination of health and wellbeing improvement metrics at the individual level and at the population level or were aimed to improve provider and managerial outcomes, in turn impacting health and wellbeing indicators. For example, an intervention aimed to revamp the electronic health records of a health centre to improve reporting and storage of health data, and early diagnosis of diabetes mellitus. This resulted in an increase in early detection and helped providers dispense better care (20).

There were certain interventions that aimed to improve quality of life, functioning, reintegration into society or improvement in social support. These resulted in the improvement of health and well-being outcomes on a population level, along with some impact on a social level. Such community based-camps or treatment programs were rare but had the potential for a large impact, such as the education and treatment camp in Kerala for patients with Lymphatic Filariasis (LF) (88). This camp provided treatment to patients and helped equip them with skills and knowledge to improve their social standing, reintegrate them into society, and educate their family members or caregivers on their condition(88). This had a multifold impact on the quality of life of these patients, social structure and awareness among people concerning this condition(88). Certain interventions were about developing protocols, and indicators for monitoring health conditions (physical and mental health), improving the capacity of primary care centres and incorporating digital aids to assist healthcare providers or continue education. These were a few that aimed to bring about changes at an organizational level. Some interventions also resulted in unintended consequences that brought about a positive change. For example, a program that was designed to reduce inappropriate prescription of antibiotics for Upper Respiratory Tract Infections (UTRI) by providers among rural communities saw not only a change in providers’ attitudes but also among the patients(89). It created greater awareness among the people to seek care earlier to be able to avoid large doses of antibiotics that may be needed in the case when such infections are left untreated (89).

## Discussion

India, given its population, diversity and economic position has been the forerunner in frugal innovations (90). Some of these innovations are not just low cost but deliver high quality care at scale. The systematic review revealed a few opportunities. Firstly, there is a lack of documentation on innovative solutions adopted by specific minority populations like people who live near large water bodies, and whose mobility is largely restricted to water, nature-based products used by tribal populations do not find mention. Several innovations mentioned seem to be localized and dependent on individual and local leadership, whereas several digital diagnostic health innovations are limited to single conditions, rendering them inefficient. In 2018, the World Health Organization declared that a people-centered health system would likely help achieve universal health coverage, rather than a health system led response. However, this is yet to be seen from the systematic review as there seems to be a lacunae in either documentation or in practice of practicing radical social participation.

The COVID-19 pandemic had a significant impact on healthcare systems worldwide, leading to the rapid adoption of innovative practices such as telemedicine, which facilitated the delivery of healthcare services to patients’ homes, reducing the risk of virus transmission while ensuring efficient care (47,51,91). Furthermore, task-shifting among healthcare providers has gained momentum during the pandemic. A significant number of these innovations were focused on non-communicable diseases (NCDs) such as diabetes, hypertension, and mental health services, while infectious diseases like tuberculosis, HIV, and COVID-19 had population-level screening innovations that utilized technology (92,93). However, few innovations focused on adapting service delivery for these diseases. This is reasonable given the severe scarcity of medical personnel in India, as highlighted in the 2019-2020 Economic Survey (94). Consequently, the majority of innovations focused on capacity building, including task shifting to community health professionals, continuous training and monitoring and incentivization. However, these innovations differed significantly by state, with southern states prioritizing health promotion programs and community ownership models over workforce restructuring initiatives prevalent in other parts of the country. Lay health workers and task shifting has been proven to be a highly effective method for outreach, but the question remains whether it is the only method of outreach that can be successful. With most of these interventions focusing on the downstream and adopting a “fix as we go” motto, it is important to look at whether these interventions are sustainable in the long run(48).

This aggregated data sheds light on multiple problematic areas along with pools of progress and innovation. With many high impact studies falling under community health ownership and lay-health worker programs, we see a large focus, and perhaps a needed one, in community integration. However, with a lack of organizational and structural/governance changes, many innovations seem to be constricted within the current system, resources and status quo, which may no longer be the most effective system for India today. We need to further understand how these innovations function within their context, what restricts them and what allows them to be successful, to understand where and what we need to prioritize when we think of furthering the primary healthcare system today. Additionally, pilot innovations are overlooked or under-resourced in the current healthcare funding landscape. Public health systems’ failure to adopt or fund these innovations is preventing their expansion at scale or their sustainability(95).

With digital interventions gaining popularity across the country, India is uniquely positioned at the intersection of technology and tradition. The adoption of digital innovations has demonstrated that combining traditionally effective public health and clinical models with new technology is very effective and can take some burden off of healthcare professionals. The successful mobile-driven community outreach program to improve maternal and neonatal outcomes by ASHA workers(48), a mobile healthcare technology to improve communication among healthcare professionals, improving patient follow-up rate and easing the workload of nurses(49), and large-scale surveillance by lay health workers for early diagnosis and record management across large populations(37), are some stellar examples of the potential of mixing digital innovations with existing ones. While digital innovations alone have not been as successful, adding a ‘human’ element has proven to be widely accepted.

While digital innovations are slowly gaining momentum, those alone cannot drive the accomplishment of universal healthcare. Multiple scalable systemic innovations are required to create salient change. Data has proven that multilayered interventions work to address many facets of the healthcare agenda. They possess the ability to address several components of the care provision integrating a holistic approach and greater impact. However, with only 10 (refer Table 5) out of a total 241 innovations that were included in the dataset, we can see that not enough priority is being given to such multilayered interventions. The current healthcare landscape of the country also demonstrates that we have been primarily focused on individual, patient-centric health outcomes. This approach, although equally necessary and valuable, misses the larger picture of what the population needs as a whole. This review highlights the gap that needs to be addressed in terms of developing interventions that target population-health, and which need major political will and backing.

The lack of preventive or early detection interventions is also pointed out by this review. The amalgamation of public health with medical science is critical to reach the most marginalized. In India, inequities in access to healthcare and poor quality of health care are significant factors underlying high levels of morbidity and mortality.

This systematic review highlighted the need for more research on developing frameworks for primary healthcare innovations in developing countries at the systemic, provider and community levels. We also identified the need for documented insights on implementation of best practices with healthcare innovations, understanding them in contexts of on-ground limitations and opportunities. To be useful to policymakers in the UHC era, health systems research must answer questions about population health, provide contextually relevant results in a format that policymakers can use, and be methodologically innovative to balance rigorous assessment of causality of outcomes with practical policy implementation considerations.

## Conclusion

This study identifies a wide range of primary healthcare innovations across different parts of India, contributing to the existing literature on primary healthcare implementation. Primary healthcare innovations play a crucial role in enhancing healthcare access, addressing health disparities, combating the disease burden, improving maternal and child health, alleviating healthcare workforce shortages, and emphasizing preventive and promotional healthcare. These innovations aim to bridge gaps by enhancing the availability, accessibility, and affordability of healthcare services, especially in underserved areas. Additionally, they target specific health needs and challenges faced by diverse communities, thereby reducing health inequities. Empowering and training community health workers is integral to these innovations, enabling them to contribute to reducing the disease burden, improving maternal and child health, and alleviating healthcare workforce shortages.

Furthermore, this review highlights a minimal number of innovations in the governance and finance categories. It is important to focus on developing innovations in these areas as they are crucial for effective primary healthcare implementation. Through the implementation of health education programs, behavior change interventions, and community engagement initiatives, primary healthcare innovations can alleviate the burden on secondary and tertiary care services, leading to overall improvements in population health. Consequently, primary healthcare innovations have the potential to revolutionize India’s healthcare landscape and foster better health outcomes for its population. The diversity of innovations among various population groups underscores the significance of contextual factors in improving primary healthcare, providing policymakers, practitioners, and researchers with valuable insights for planning and developing future interventions towards achieving universal health coverage.

## Data Availability

The data described in systematic review is openly available in Primary Healthcare Innovations - https://www.learning4impact.org/phc-innovations

## Supporting information - Funding

This systematic review was co-funded by the Health System Transformation Platform(HSTP) under the PHC Landscape Study project, USAID under the Learning4Impact project and Swasti. All parties were involved in the study design, interpretation, and submission decisions for publication.

## Acknowledgments

The authors would like to acknowledge the contributions Shiv Kumar, co-founder Catalyst Group, Swasti and Dr Rajeev Sadanandan, CEO, Health Systems Transformation Platform, Tata Trusts. A mention also for the colleagues who supported us in this review namely –Sabhimanvi Dua, Meghana Ratna Pydi, Purnima Ranawat, Ajinkya Mujumdar, Ankita Elizabeth Mathew, Govind Gopalan Nair, Mikala Kowal, Namita Varma, Uneet Kumar Singh.

## Authors and Affiliations

CEO, Swasti Health Catalysts, Bangalore, India Angela Chaudhuri

Chair TB PPM Learning Network, India Chapter, McGill University, Canada Formerly in Health Systems Transformation Platform Vijayshree Yellappa

Health Research Lead, Swasti Health Catalysts, Bangalore, India Neha Parikh

Research Associate, Swasti Health Catalysts, Bangalore, India Ranjana N Rao

Technical Specialist, Swasti Health Catalysts, Bangalore, India Nilakshi Biswas

Primary Healthcare Exemplars Consultant, Swasti Health Catalysts, Bangalore, India Nandini Agarwal

Practice Manager - Health Consulting,Swasti Health Catalysts, Bangalore, India Catherine Cove

Public Health Specialist, Swasti Health Catalysts, Bangalore, India Bhumika Nanda

## Author Contributions

AC and VY conceptualized the research. All the authors (AC,VY,NP,NB,NA,BN,CC,RNR) screened the titles and abstracts of all papers and the full text of all articles remaining were obtained and reviewed. All Authors participated in developing study methods, definitions and criteria. AC and VY participated in the sequence in drafting the manuscript.VY, AC, NP, RNR edited and reviewed the final manuscript.

**Note:**The authors - AC and VY have equally contributed and both authors have mutually agreed to share 1st authorship.

